# Accuracy verification of low-cost CO_2_ concentration measuring devices for general use as a countermeasure against COVID-19

**DOI:** 10.1101/2021.07.30.21261265

**Authors:** Yo Ishigaki, Koji Enoki, Shinji Yokogawa

## Abstract

Within the context of the COVID-19 pandemic, CO_2_ sensors that measure ventilation conditions and thereby reduce the risk of airborne infection, are gaining increasing attention. We investigated and verified the accuracy of 12 relatively low-cost sensor models that retail for less than $45 and are advertised as infection control measures on a major e-commerce site. Our results indicate that 25% of the tested sensors can be used to identify trends in CO_2_ concentration, if correctly calibrated. However, 67% of sensors did not respond to the presence of CO_2_, which suggests that a type of pseudo-technique is used to display the CO_2_ concentration. We recommend that these sensors are not suitable for infection prevention purposes. We also found that all 67% of the sensors that did not respond to CO2 responded strongly to alcohol. Owing to the widespread use of alcohol in preventing the spread of infectious diseases, sensors that react to alcohol can display inaccurate values, resulting in inappropriate ventilation behavior. Therefore, we strongly recommended that these sensors not be used. Based on our results, we offer practical recommendations to the average consumer, who does not have special measuring equipment, on how to identify inaccurate CO_2_ sensors.

## Introduction

In enclosed spaces with inadequate ventilation and air treatment, the airborne spread of SARS-CoV-2 can occur even at a distance of 6 feet or more from an infected person, especially as concentrations of very fine droplets and aerosol particles drift through the air [1]. Within this context, airborne transmission may have accounted for more than 50% of the disease transmission on the Diamond Princess, the cruise ship associated with the SARS-CoV-2 outbreak in Japan [2].

The concentration of indoor carbon dioxide (CO_2_) has attracted attention as a proxy for infection risk [3]. While outdoor fresh air has a constant CO_2_ concentration of approximately 400 ppm, human breath contains a much larger amount of CO_2_ at 30,000–40,000 ppm. Therefore, by measuring indoor CO_2_ concentration, it is possible to understand how much human breath is retained in the air.

The American Society of Health Engineers (ASHE) states that there is a correlation between ventilation capacity and the risk of airborne infection from tuberculosis, measles, chicken pox, influenza, smallpox, and SARS [4]. The Wells-Riley equation (Equation 1) can be used to quantitatively evaluate the risk of airborne infection by bacteria and viruses [5-6].

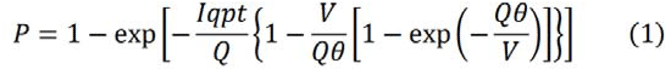

In this equation, *P* is the probability that an infected person will be infected, *I* is the number of infected persons in the enclosed space, *p* is the respiratory volume of one person (m^3^/h), *q* is the rate of infectious droplets (/h), *t* is the time spent by the infected person in the space (h), *θ* is the time spent by the infected person in the space (h), *V* is the volume of the room (m^3^), and *Q* is the ventilation rate (m^3^/h). Thus, the Wells-Riley equation models the probability of exposure to infectious droplets emitted by an infected person and inhaled by an uninfected person through respiration as the risk quantity. As infectious droplets are accompanied by a high concentration of CO_2_, both of which are airborne, it is beneficial to use CO_2_ concentration as an indicator of the airborne infection risk.

A number of researchers have suggested that the threshold ventilation rate for maintaining the effective reproduction rate of tuberculosis (the ratio of new secondary infected persons to the source of infection) below 1 corresponds to a CO_2_ concentration of 1,000 ppm. Specifically, Du et al. found that by improving the ventilation system in a poorly ventilated university building, where an outbreak of tuberculosis occurred (27 tuberculosis patients and 1,665 contacts), and by reducing the maximum CO_2_ concentration from 3204 ± 50 to 591-603 ppm, the secondary infection rate among new contacts could be reduced to zero (mean follow-up, 5.9 years). Furthermore, the incidence of tuberculosis among contacts could be reduced by 97% (95% CI: 50–99.9%) when the CO_2_ concentration remained below 1,000 ppm [7].

After comprehensively considering laws and regulations concerning CO_2_ concentration, the Occupational Hygiene and Ergonomics Subcommittee of the Japan Society for Occupational Health (JSOH) established classifications for estimating ventilation status according to CO_2_ concentrations (Table 1), and recommended measures according to each classification [8].

**Table 1.**
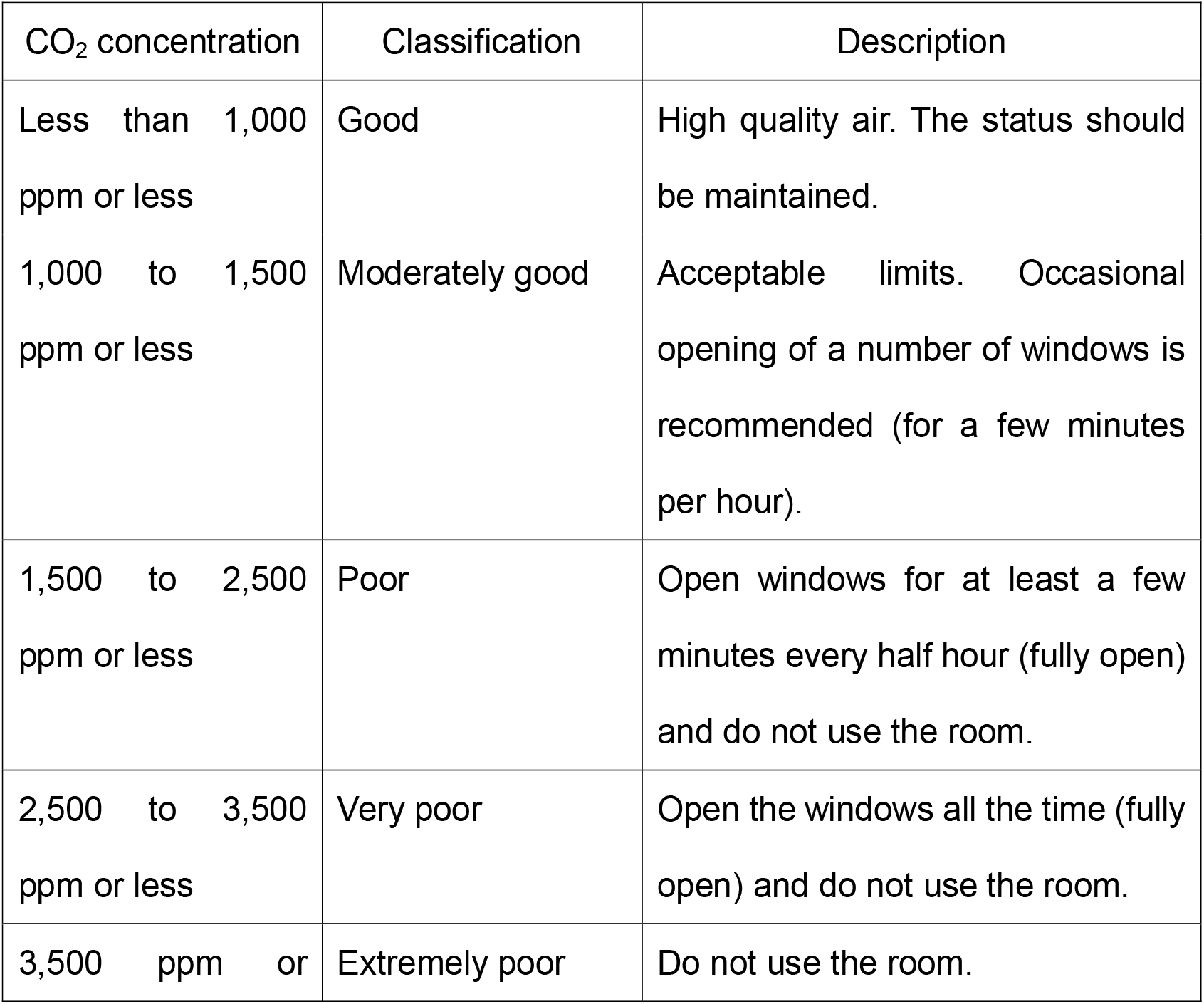
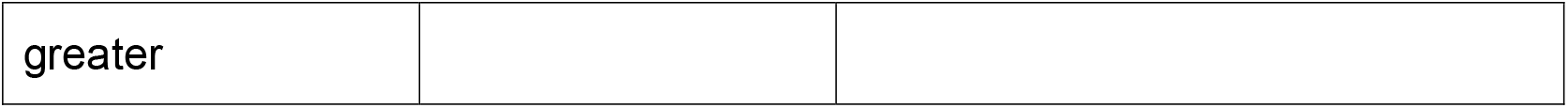
Ventilation criteria based on CO_2_ concentrations, established by The Occupational Hygiene and Ergonomics Subcommittee of the Japan Society for Occupational Health (JSOH).

Like those of other countries, the Japanese Building Standard Law (revised and implemented in 2003), requires the installation of a 24-hour ventilation system in all living rooms to prevent the development of the sick building syndrome, and requires an air change rate per hour (ACPH) of 0.5 times/h or more inside houses. With an ACPH of 0.5 times/h, for example, in a 70 m^2^ space with a standard activity level, the CO_2_ concentration at saturation is 800 ppm with two people in the room, 1,000 ppm with three people in the room, and 1,200 ppm with four people in the room.

While the Building Standard Law specifies the amount of ventilation, Japan’s Building Management Law (enacted in 1970) specifies CO_2_ concentration standards. These laws specify that in buildings used as entertainment venues, department stores, restaurants, shops, offices, and schools, where the enclosed area used for specific purposes is 3,000 m^2^ or more, defined as “specified buildings” (> 8,000 m^2^ for schools), the CO_2_ concentration should be 1,000 ppm or less.

Against this background, the monitoring of CO_2_ concentration by sensors is currently being advocated to reduce the risk of COVID-19 in indoor environments. In many instances, especially in the context of a pandemics, it is recommended to maintain indoor CO_2_ concentrations below 800 ppm (e.g., Minnesota Department of Health, Federation of European Heating Ventilation and Air Conditioning Associations, UK Scientific Advisory Group for Emergency, US Centers for Disease Control and Prevention) [9]. In Japan, a widespread system is used, in which local government supports the cost of installing CO_2_ sensors in restaurants to prevent the spread of COVID-19. For example, the Tokyo Metropolitan Government, Kyoto Prefecture, and Osaka City support the purchase of infection prevention devices, including CO_2_ sensors, up to 100,000–300,000 yen. In addition, Shibuya Ward in Tokyo distributes free CO_2_ sensors to restaurants located in the ward.

In response to this sudden increase in demand, low-cost CO_2_ sensors have rapidly become available on e-commerce sites such as Amazon, and are advertised as being effective in preventing the spread of infectious diseases. Many of these sensors have vague descriptions of their measurement principles, with several consumers questioning the CO_2_ measuring ability of these sensors. In addition, there is a lack of guidelines for the use of CO_2_ sensors in Japan, and a lack of laws or regulations regarding their accuracy and reliability. In response to this research need, we purchased several low-cost CO_2_ sensors, to verify their accuracy and investigate the related measurement principles.

Although several studies have already evaluated low-cost gas sensors [10, 11], including CO_2_ sensors, the global outbreak of COVID-19 has led to a proliferation of new products on the market for the control of infectious diseases that have not been tested for efficacy and accuracy. Therefore, we conducted this study to investigate the accuracy and reliability of these sensors and to offer advice on these sensors to the general consumer, who does not have access to specialized inspection equipment.

## Methodology

In this study, we purchased and investigated 12 relatively low-cost CO_2_ sensors (1–12; Table 2), within a price range of 2,900–4,999 JPY (approximately 26–45 USD, at the exchange rate in July 28, 2021) from the Amazon Japan website, and installed them according to instructions. The sensors were advertised as being effective in preventing COVID-19 infections. In addition to the 12 sensors, we purchased two relatively expensive CO_2_ sensors, (A and B) that are widely used for industrial and experimental purposes as reference devices. Sensor A is manufactured by the T&D corporation TR-76Ui (54,780 JPY, approximately 500 USD), with a zero and 1500 ppm span calibration, while Sensor B is manufactured and factory calibrated by the C.H.C. System corporation NMA-PR-R (47,500 JPY, approximately 430 USD). Both sensors A and B are based on the non-dispersive infrared (NDIR) method.

**Table 2.**
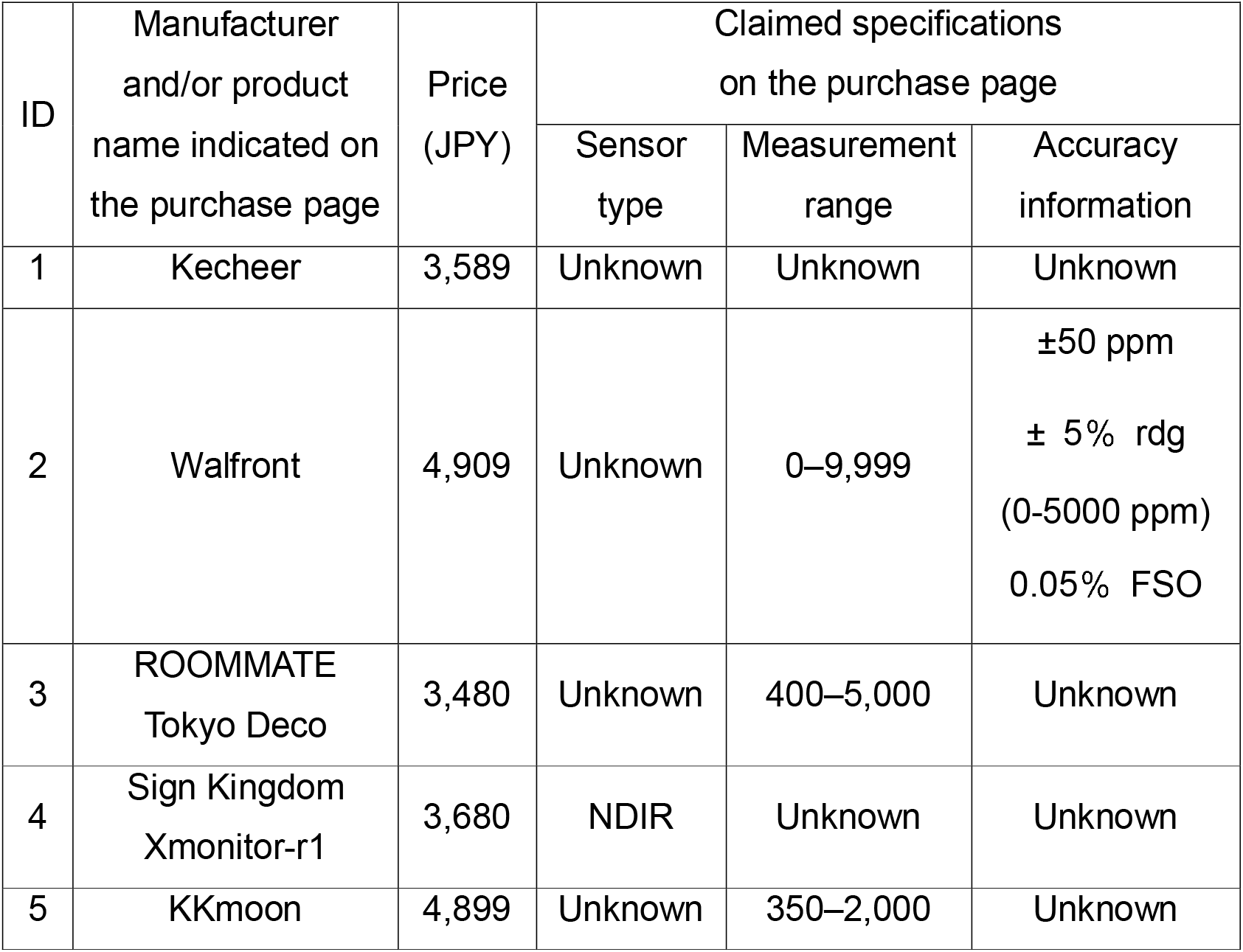
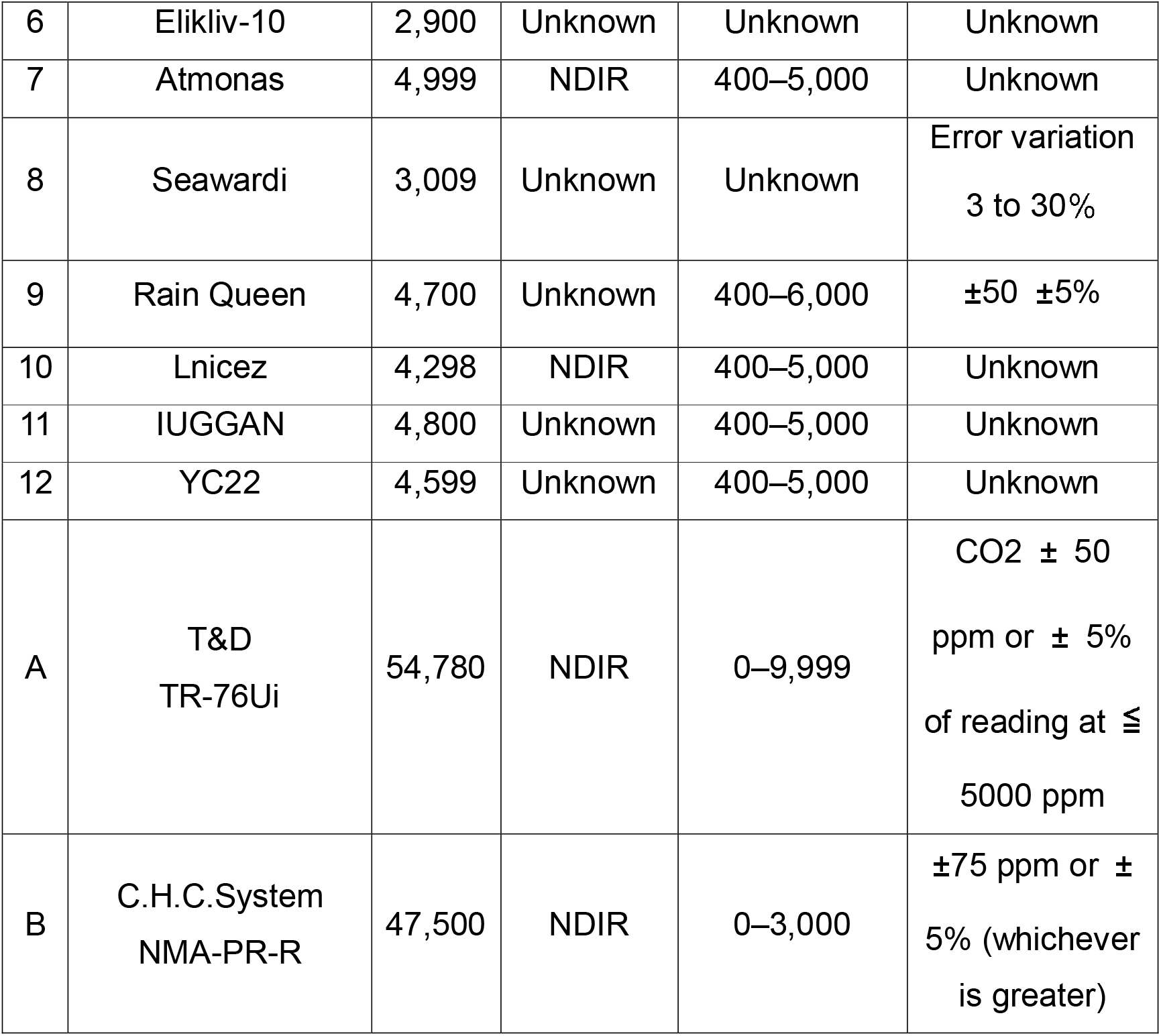
List of CO_2_ sensors used in the experiment.

All 14 sensors (1 to 12, A and B) were activated simultaneously and placed in a chamber (Fig. 1). The following protocol was used for the experiment:

**Figure 1.**
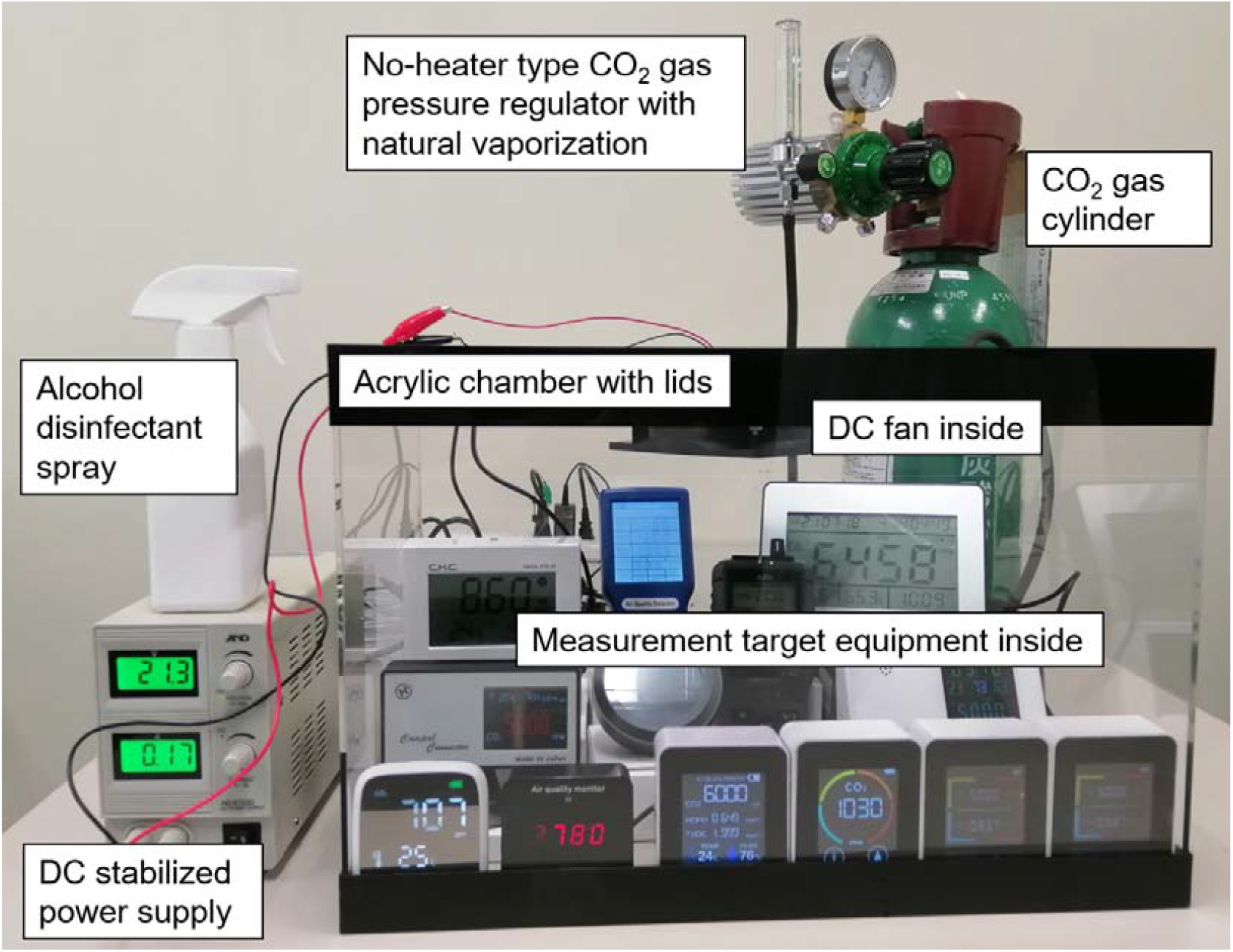
Image of experimental equipment and setup used for an investigation of the accuracy of 12 CO_2_ sensors.

1. Open the chamber lid.
2. Turn on the fan.
3. Ventilate the chamber with room air (adjusted to 25°C) for 5 min.
4. Close the chamber lid.
5. Turn off the fan.
6. <FRESH AIR> Record the displayed values of each measuring instrument. However, if a value outside the measurement range is displayed, it is not recorded.
7. Turn on the fan.
8. <INJECTION> Inject (additional) CO_2_ gas, (additional) room air and/or alcohol into the chamber to change the gas concentration in the chamber.
9. Wait for 5 min until the inner air and gas are evenly mixed in the chamber.
10. Turn off the fan.
11. <CONTROLLED ATMOSPHERE> Record the displayed values of each measuring instrument. However, if a value outside the measurement range is displayed, it is not recorded.
12. Return to step 7 (or terminate the experiment).

## Results and Discussion

We compared the CO_2_ values of the 12 investigated sensors to those of the reference sensors (A and B; Fig. 2). The relatively expensive sensor B showed a narrower measurement range than sensor A, although both had very similar response characteristics. Sensor 10 generally displayed higher values than sensor A, while sensors 4 and 7 displayed lower values than sensor A. Although sensor 2 responded to CO_2_, it seemed to malfunction, as it displayed very high CO_2_ values. Sensors 1, 3, 5, 6, 8, 9, 11 and 12 did not seem to respond to CO_2_ levels. As these eight sensors are able to display total volatile organic compound (tVOC) values, we conducted an additional experiment by applying approximately 5 ml of rubbing alcohol to the inside of the acrylic plate of the chamber.

**Figure 2.**
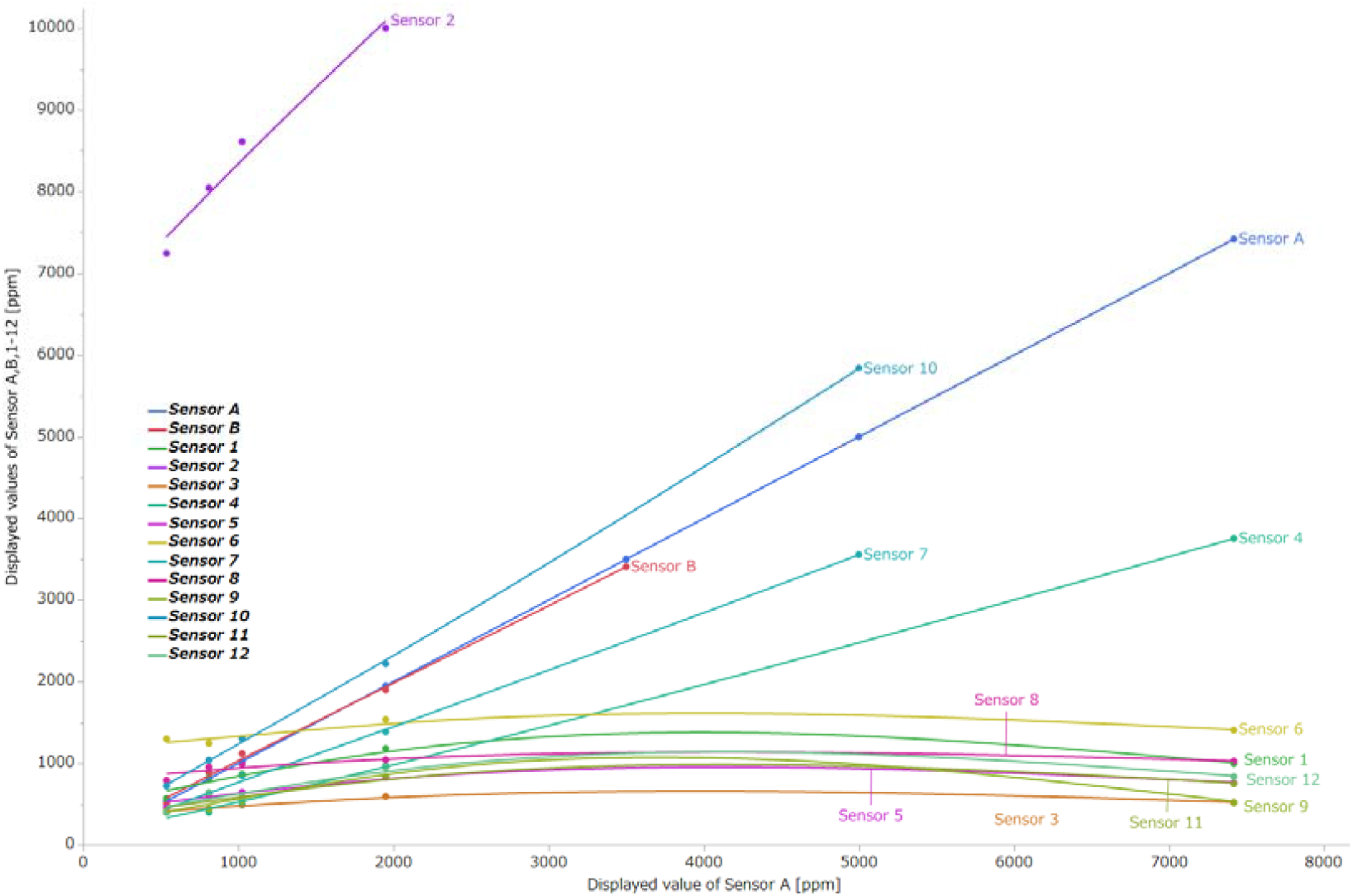
Response characteristics of CO_2_ sensors. Measured values of sensor A, the reference device, are plotted on the horizontal axis, and the measured values of sensors A, B, and 1 to 12 are plotted on the vertical axis, connected by spline curves.

During this experiment, rather than adding CO_2_ gas, 5 ml of alcohol was sprayed on the acrylic wall in the chamber (Fig. 3). To eliminate the cross-recruitment and contamination effect of alcohol, the experiment was performed independently to the previous experiment.

**Figure 3.**
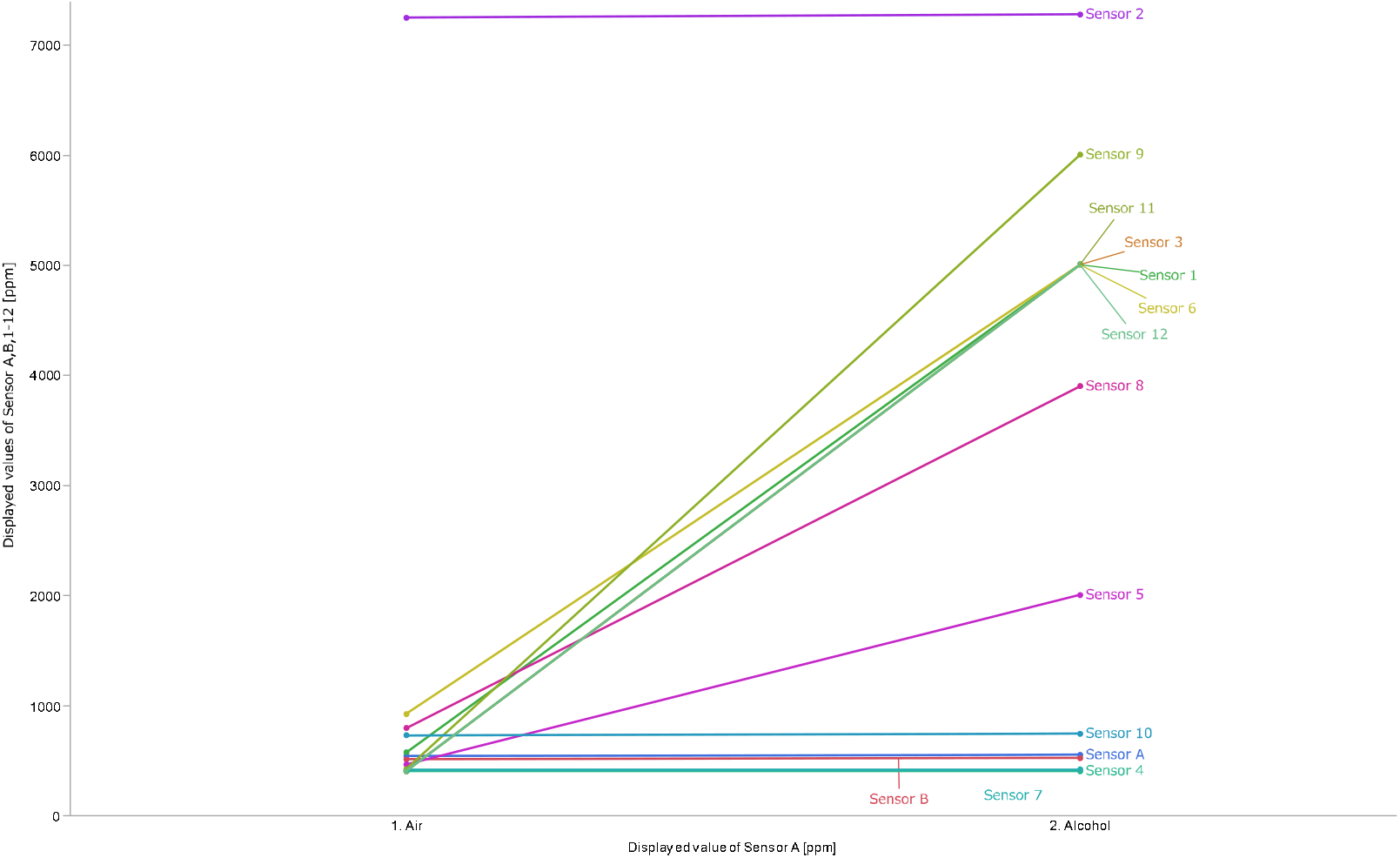
Response characteristics of 14 CO_2_ sensors to alcohol, including two reference sensors (A and B). “1. Air” on the x-axis corresponds to the sixth step (FRESH AIR) in the experimental protocol, while “2. Alcohol” refers to the twelfth step of the protocol (CONTROLLED AIR), with 5 ml of alcohol added to the chamber at the step 8 (INJECTION) instead of CO_2_.

Our results (Fig. 3) indicate that the readings of sensors A, B, 2, 4, 7, and 10 were not affected by the presence of alcohol. On the other hand, sensors 1, 3, 5, 6, 8, 9, 11 and 12 reacted strongly to alcohol. Furthermore, these eight sensors match the sensors that did not respond to CO_2_ (Fig. 2).

Of the 12 relatively low-cost sensors investigated in this study, three models (sensors 4, 7, and 10; 25% of the total) can potentially be used to determine CO_2_ concentration levels, if correctly calibrated. However, the accuracy of the sensors was not as good as that of the more expensive reference sensors. Machine learning methods have been proposed for calibration of inexpensive sensors [12, 13]. Sensor 2 seemed to be malfunctioning (8% of the total), while eight sensor models (67%) did not respond to CO_2_, and they responded strongly to alcohol, suggesting that the sensors were equipped with different sensors to the CO_2_ sensing module.

To investigate the sensing module, we purchased and disassembled an extra model of sensor 12 (Fig. 4). Notably, the NDIR sensor was not present. Based on the model number of the sensing module and the manufacturer’s specifications [14], we identified that the sensor was not designed for CO_2_ measurements, but rather for the measurement of “ammonia, hydrogen, alcohol, carbon monoxide, methane, propane, Gan, styrene, propylene glycol, alkyl phenol, toluene, ethylbenzene, xylene, formaldehyde, and other volatile organic gases, incense smoke, wood, paper smoke burning out.” This suggests that sensor 12 is designed to display the concentration of CO_2_ by a type of imitating algorithm based on the value of the sensing module that detects miscellaneous gases, excluding CO_2_.

**Figure 4.**
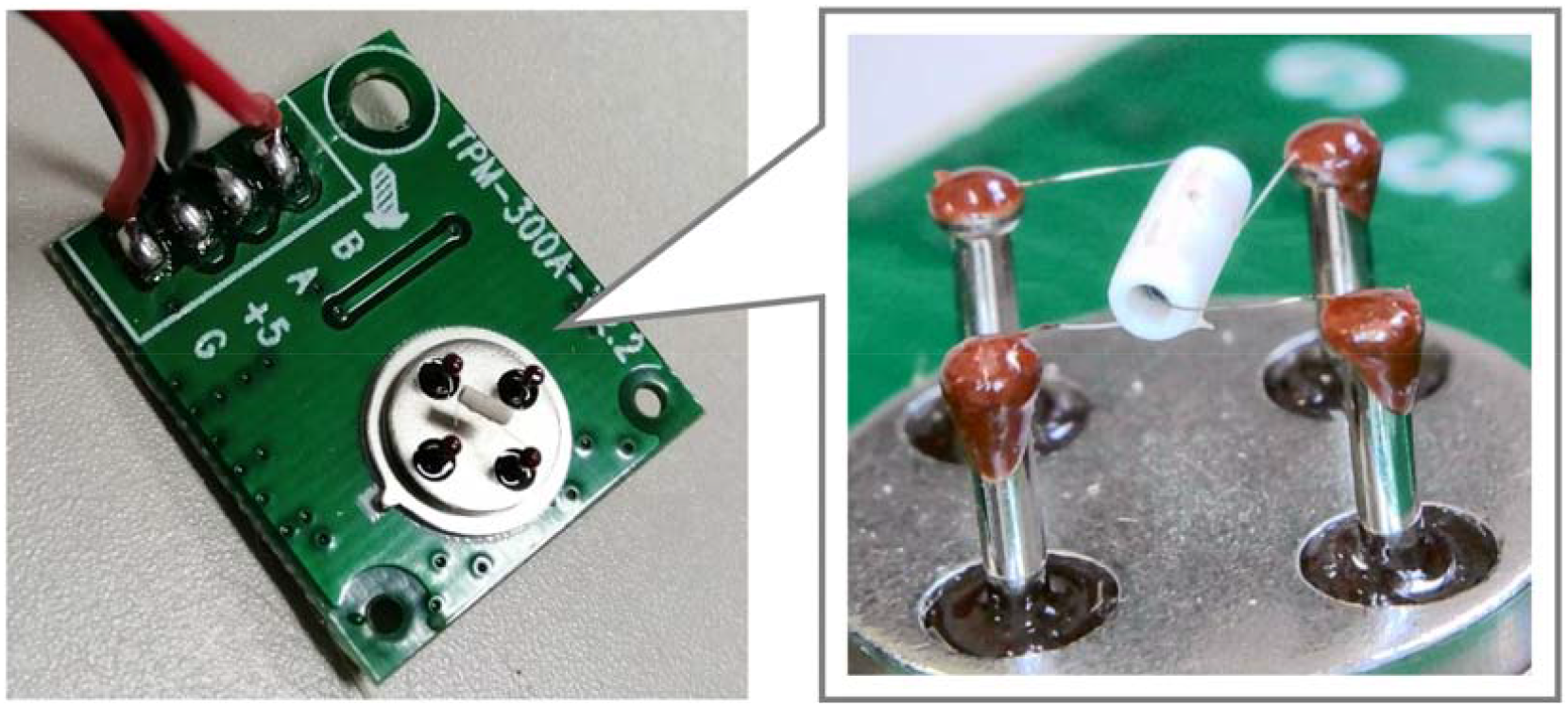
Sensing module of Sensor 12. A cylindrical object was suspended in the air by four wire bonds, and inside it was a coil heater.

Globally, several sensors display a very vaguely defined measurement called eCO_2_ (equivalent CO_2_). This is estimated from the tVOC value and has been shown to differ from the actual CO_2_ concentration [15].

Sensors 1, 3, 6, 8, 9, 11, and 12, which responded strongly to alcohol (Fig. 3), all displayed tVOC values separately from the CO_2_ concentration. Therefore, we suspected that these six sensor models were displaying eCO_2_ values rather than actual CO_2_ concentrations. However, the sales pages for these six sensors did not indicate that they used eCO_2_ sensors.

Based on the results of our experiments, we developed a simple method for consumers to verify the quality of sensors on offer, without needing specialized equipment. Within this context, we recommend the following:

1. Ensure that the sensor displays a value of approximately 400 ppm in fresh air outdoors (this will allow for the detection of faulty sensors such as sensor 2, with a large deviation in CO_2_ values).
2. Ensure that the CO_2_ concentration rises significantly when breathing on the sensor (this will help identify sensors that do not respond to increases in CO_2_, such as Sensors 1, 3, 5, 6, 8, 9, 11 and 12).
3. Ensure that the sensor does not react erroneously to alcohol by holding a hand dipped in an alcohol disinfectant close to the sensor (this eliminates low-quality sensors that pseudo-indicate CO_2_ concentration from tVOC concentrations such as alcohol).

## Conclusions

Twenty-five percent of the relatively low-cost sensors investigated in this study have the potential to be used for identifying trends in CO_2_ concentration, if correctly calibrated, although with a poor accuracy. Notably, 67% of the sensors did not respond to changes in CO_2_ levels, which suggests that various pseudo-masquerading techniques are being used to display CO_2_ concentrations. These sensors should not be used to prevent the spread of infectious diseases. In addition, the 67% of the sensors reacted strongly to alcohol. As alcohol is widely used indoors to prevent the spread of COVID-19, sensors that respond to alcohol will display inflated values, which can lead to incorrect ventilation behavior. Therefore, we strongly recommended that these sensors not be used.

This study was conducted using a total of 14 commercially available sensor models. However, there are many models of CO2 sensors available in the market. We suggest that future research focus on an exhaustive analysis of the sensors available in the market as a basis for a discussion on the guidelines and regulations for CO_2_ sensors. We are planning to use gas chromatography with an accuracy of ±0.1 ppm to simultaneously detect the concentration of alcohol and carbon dioxide for more accurate verification. With this method, we will be able to measure both organic and inorganic gases, which was not possible before.

In addition, clarification of the electrical circuit, catalyst, semiconductor composition, and solid-state properties of the sensing modules (Fig. 4) will assist to pinpointing the purpose for which these sensors were developed. Furthermore, we suggest that future research focus on not only conducting exposure experiments with pure CO_2_ gas, but also include accurate measurements with human-derived CO_2_ gases, including various volatile organic compounds.

## Data Availability

The raw data of the graphs are available on FigShare.

https://figshare.com/articles/figure/Accuracy_verification_of_low-cost_CO2_concentration_measuring_devices_for_general_use_as_a_countermeasure_against_infectious_diseases/15067557

## Notes

The authors declare no conflicts of interest. This paper has been previously submitted to medRxiv, a preprint server for health sciences. This study was approved by the Ethics Committee on Experiments on Human Subjects (approval number of 21005), The University of Electro-communications, located at Chofugaoka 1-5-1, Chofu, Tokyo, Japan.

## Acknowledgments

This work was supported by the Research Grant Program of the KDDI Foundation. In addition, some of the data acquisition technologies in this study are based on results obtained from a project commissioned by the New Energy and Industrial Technology Development Organization (NEDO).

## Data availability

The raw data of the graphs are available on FigShare.

